# GOOGLE TRENDS IN THE DIAGNOSIS OF METEOSENSITIVITY

**DOI:** 10.1101/2024.11.15.24317370

**Authors:** Ksenia Yarovaya, Iryna Hnatiuk, Yuliia Solodovnikova, Anatoliy Son

## Abstract

**Introduction:** Primary headache disorders, including tension-type headaches (TTH) and migraines, are highly prevalent and pose a significant socio-economic burden. Environmental triggers, such as weather changes, are commonly believed to provoke these headaches.

**Methods:** This study aims to explore the role of geomagnetic activity as a potential trigger for TTH and migraines. Using Google Trends data, we tracked public interest in the search terms “migraine” and “headache” in Ukraine from 2019 to 2023. We analyzed the Ap index, which measures geomagnetic activity, alongside the search data to explore potential correlations.

**Results:** A total of 261 observations were made. The analysis revealed a negative correlation between the popularity of search queries for “migraine” and “headache” and geomagnetic activity, with increased search interest during periods of low geomagnetic activity. TTH patients were found to be more sensitive to low geomagnetic activity compared to those with migraines, suggesting different pathophysiological mechanisms between these two types of headaches. While the correlation is statistically significant, it remains relatively weak, indicating that geomagnetic fluctuations may contribute to headache triggers but are not the sole factor.

**Conclusions:** The findings suggest a potential link between geomagnetic activity and the incidence of primary headaches, with TTH being more responsive to these fluctuations than migraines. The study underscores the need for further research into the impact of geomagnetic activity on human health and headache disorders.

**What is already known on this topic:** Primary headache disorders, such as tension-type headaches (TTH) and migraines, are commonly triggered by environmental factors, but the role of geomagnetic activity remains underexplored despite its known effects on human health, like circadian rhythms and blood pressure.

**What this study adds:** This study identifies a significant negative correlation between low geomagnetic activity and increased public interest in headache-related terms, suggesting that geomagnetic fluctuations may act as environmental triggers for headaches, particularly TTH. It highlights distinct sensitivities between TTH and migraines to geomagnetic changes and demonstrates the utility of Google Trends as a tool for exploring public health patterns related to environmental factors.

**How this study might affect research, practice or policy:** This study highlights geomagnetic activity as a potential environmental trigger for primary headaches, encouraging further research into its role and distinct impacts on TTH and migraines. It underscores the utility of tools like Google Trends for public health surveillance, offering insights for healthcare practices and policy development to better address headache prevention.

## Introduction

Primary headache disorders, including tension-type headache (TTH) and migraine, have a wide prevalence, occurring in nearly everyone and imposing a significant secondary socio-economic burden due to the impact on the working population [1]. The trigger for headaches is any factor that independently or in combination with others can initiate or contribute to the neurobiological process leading to headaches. A trigger can occur in close temporal proximity to the headache phase and usually occurs less than 3 hours before an attack, but can occur up to 3 days before the headache phase [2]. Established triggers for migraine attacks include stress, menstruation, visual stimuli, weather changes, nitrates, fasting, consumption of tyramine-containing foods, etc [3]. The most common triggers for TTH are stress, sleep deprivation, and psychological stress [3].

Among patients, there is a widespread belief that changes in weather conditions significantly contribute to the provocation of primary headaches [4]. Based on population-based diary studies, the majority of patients suffering from migraine associate weather changes with the development of headache attacks [5,6]. Other observations suggest that weather is a trigger for tension-type headaches compared to patients with other types of headaches [7]. However, to date, environmental factors as triggers for provoking primary headaches are not sufficiently convincing [8-10].

It is well known that various manifestations of space weather can affect a wide range of human activities, from technological systems to human health. Several scientific studies show that the cardiovascular, nervous, and other functional systems react to changes in geophysical factors [11]. The presence of electromagnetic activity is associated with the existence of the magnetosphere - a region of space around the Earth where the Earth’s magnetic field dominates. Processes in the magnetosphere are very dynamic, which determines the existence of “space weather”, the change of which can affect communication systems, navigation, and human health. Geomagnetic indices A are used to measure Earth’s geomagnetic activity. Indices A are based on measurements of the horizontal components of various magnetic stations. The A index for polar latitudes (Afr) represents the average amplitude values of fluctuations in the horizontal component of the magnetic field over the last hour in the range from 60 to 75 degrees of northern and southern latitudes. The higher the value of the A index for polar latitudes, the stronger the geomagnetic activity in these regions. The planetary A index (Ap) is the planetary average value of the A index calculated taking into account measurements from several magnetic stations around the Earth. Predicted and observational values of A indices are used to determine the current state and future changes in geomagnetic activity on the Earth’s surface [12].

The results of previous research indicate a negative impact of extreme geomagnetic storms on the functional state of the human brain [11]. This disrupts the normal functioning of integrative nonspecific systems located within the limbic-reticular complex and is responsible for creating an appropriate level of arousal. Additionally, there is an imbalance of activating and deactivating mechanisms, including dysfunction of ergo- and trophotropic suprasegmental centers. The threshold of seizure readiness of the human brain decreases, which is particularly dangerous for high-risk individuals [11]. Changes in human physiological parameters (heart rate, RR interval, systolic and diastolic blood pressure) are associated with geomagnetic disturbances and variations in cosmic radiation intensity. Physiological parameters of humans demonstrated peak values (increases and decreases) for high geomagnetic activity, as well as for the greatest decrease in cosmic radiation intensity. Various types of cardiac arrhythmias are associated with different solar parameters and geomagnetic activity parameters (solar flares, sunspot number, etc.) and cosmic radiation intensity for both the 22nd and 23rd solar cycles. Patients with ventricular single and multiple extrasystoles are more sensitive to changes in solar magnetic field polarity compared to patients with supraventricular extrasystoles and supraventricular paroxysmal tachycardia [13]. Analysis of 788 cases of sudden cardiac death shows that during periods of low geomagnetic activity and on days of high-intensity geomagnetic storms, as well as the day following them, the number of sudden cardiac death cases increases [14]. Analysis of data from the Kaunas Stroke Register from 1986 to 2010 demonstrated that an increased risk of stroke may be associated with geomagnetic storms, low geomagnetic activity, stronger solar flares, and solar proton events [15]. A single-center retrospective observational study concluded that while weather conditions, including temperature, atmospheric pressure, humidity, rainfall, sunshine, and air pollutants, showed no correlation with the risk of febrile seizures in children [16]. Lower daily maximum temperatures are significantly associated with an increased risk of hospital admissions for hypertension, while acutely changing weather conditions, such as fluctuations in temperature, atmospheric pressure, and precipitation over two to three days, do not significantly impact the risk of hypertensive episodes, contrary to the perceptions of many hypertensive patients [17]. Investigating the effects of atmospheric pressure changes on intracranial pressure (ICP) in hydrocephalus patients, it was hypothesized that variations in atmospheric pressure influence arterial CO2 partial pressure, which then affects intracranial blood volume and ICP; findings suggest that a 50 hPa change in atmospheric pressure could result in a significant ICP variation of over 1,65 mmHg, potentially explaining the symptoms reported by patients [18]. The study using data from over 18,000 participants in the Tromsø Study 7 found that weather has a causal and dynamic effect on pain tolerance, with cold pain tolerance showing clear seasonal variation and correlating with meteorological variables, while pressure pain tolerance demonstrated short-term fluctuations similar to meteorological timescales [19]. Among 133 participants in a Japanese hospital survey, 32% reported physical symptoms related to bad weather, with headaches being the most common complaint, and those affected were found to be younger and more likely to experience irritability, gastrointestinal issues, and sleep disorders [20]. Extreme weather events are significantly associated with increased risks of asthma-related outcomes, including exacerbations, hospital admissions, and mortality, with heightened risks observed in children and females [21]. Evaluating data from 1,845 Finnish adults, higher average solar insolation over the year was not linked to the total number of depressive symptoms but was associated with a reduced likelihood of suicidal thoughts and an increased likelihood of changes in appetite, sleep, and feelings of worthlessness or guilt, indicating that sunlight may influence specific depressive symptoms [22]. A retrospective analysis of 742 patients from the Breton Regional Observatory on Myocardial Infarction registry revealed that in Brest, France, the incidence of ST-elevation myocardial infarction was significantly associated with a decrease in air temperature, an increase in atmospheric pressure, and higher humidity over the preceding 7 days [23]. The study found that temperature fluctuations and interactions between temperature and relative humidity significantly increased the likelihood of multiple sclerosis clinic visits among US veterans, highlighting the impact of meteorological conditions on multiple sclerosis symptom exacerbations [24]. In a study of 98 adults with episodic migraine, higher relative humidity increased migraine risk during the warm season, while traffic-related air pollutants were linked to higher migraine risk during the cold season [25]. Exposure to hot or cold weather did not significantly impact the clinical features of migraines or tension-type headaches, although, in obese patients with tension-type headaches, hot weather was more likely to trigger headaches [26]. Women are nearly three times more likely than men to experience headaches triggered by low barometric pressure, pointing to the need for sex-specific therapeutic approaches [27]. The results of another study indicated that geomagnetic disturbances, including the level of geomagnetic activity, the number of sunspots, and high proton flux events may influence the activity of systemic lupus erythematosus. Additionally, an increase in the geomagnetic activity index Ap and high-energy proton flux was associated with decreased activity of the systemic lupus erythematosus [28]. In contrast, an increase in the sunspot number index predicted a decrease in systemic lupus erythematosus activity [28].

The objective of this study is to identify environmental factors that may act as triggers for migraines and tension-type headaches (TTH).

## Materials and methods

Google Trends is a free, open-source tool used to track public interest in search terms entered into the Google search engine. Analysis with Google Trends can be configured based on search terms, period, and geographic location. After entering a search term and setting the corresponding time and geographic constraints, Google Trends generates visual elements and results that depict the volume of the search term relative to its peak popularity over a specified time. Data is presented as relative search volume, calculated as the percentage of searches for the term in a location during a specific time. A relative search volume of 100 indicates the highest proportion of searches for a particular topic compared to the total number of Google queries. A value of 0 indicates that at the specified moment, the share of searches for the search term was less than 1% of its maximum relative search volume [29].

Data on search queries for “headache” and “migraine” are obtained from Google in Ukraine weekly from 2019 to 2023. The dataset consists of 261 observations. The obtained indicators reflect the relative popularity of the query, formed by the ratio of the selected query to the total number of queries and expressed on a scale of 100. Since TTH is the most prevalent primary headache and shares several nonspecific clinical characteristics, in further analysis, the results for the query “headache” are considered indicative of interest in TTH.

To determine the state and changes in geomagnetic activity on the Earth’s surface, Ap data from the archives of the Space Weather Prediction Center of the National Centers for Environmental Information are used. Ap values close to 0 indicate low geomagnetic activity, while values of Ap close to 100 and above indicate high activity [30].

## Results

The popularity of public interest in the Google search engine can be an important tool for understanding and predicting migraine and TTH triggers. The relative volume of searches in Google Trends is geographically concentrated on the territory of Ukraine (Graph 1).

**Graph 1:**
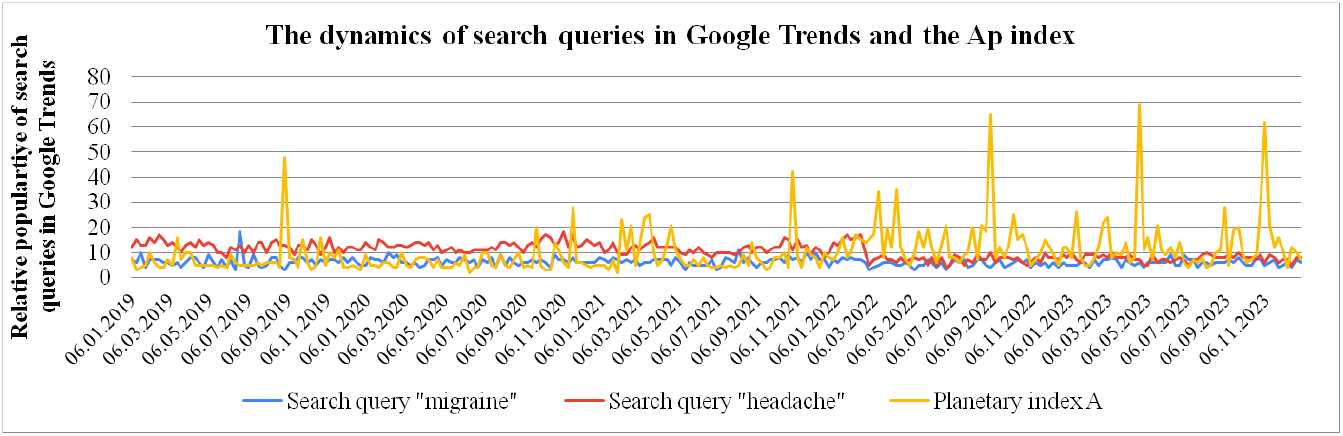
Dynamics of search query indicators in Google Trends and the Ap index.

The database analysis covers the period from 2019 to 2023. The Ap indicator is used to determine the relevance of users’ anxiety about TTH and migraine and geomagnetic storms as triggers of primary headaches. The demand for information on migraines and headaches indicates the significance of these conditions in Ukraine. In this study, an analysis of descriptive statistical indicators (Table 1) is conducted to examine the relationship between the occurrence of TTH and migraine with values of the geomagnetic index Ap.

**Table 1.** Summary statistics for search queries (“migraine” and “headache”) and Ap index during the period 2019–2023.

|  | The search query<br>“migraine” | The search query<br>“headache” | Ap |
| --- | --- | --- | --- |
| N | 261 | 261 | 261 |
| Average | 6,20 | 10,5 | 10 |
| SD | 1,62 | 3 | 8,95 |
| Median | 6 | 10 | 8 |
| Minimum | 3 | 4 | 2 |
| Maximum | 18 | 18 | 69 |

The average value of the search term “migraine” is 6.20 ± 1.62, while the average value of the search term “headache” is 10.5 ± 3, indicating significantly higher interest and a greater spread of popularity for queries related to “headache” compared to “migraines”. The median for the search terms “migraine” and “headache” is 6 and 10 respectively. The minimum value in the dataset for the “migraine” query is 3, and for the “headache” query is 4. The maximum popularity of the “migraine” and “headache” queries were the same, equaling 18 in both cases. When analyzing space weather, the average level of Ap during the study period is 10 ± 8.95, indicating significant fluctuations in the Earth’s magnetic field. The median of the Ap index is 8. Extreme variations in geomagnetic activity based on the Ap index range from 2 to 69. A total of 261 observations were obtained for each category over the study period.

The results of the correlation analysis (Table 2) indicate a negative correlation between the popularity of searches for “migraine” and “headache” on Google and changes in geomagnetic activity measured using the planetary index Ap in Ukraine.

**Table 2.** Correlation analysis data.

| Correlation analysis |  |  |  |
| --- | --- | --- | --- |
|  | r | df | p-value |
| Search query “migraine” and Ap | -0,123 | 259 | 0,048 |
| Search query “headache” and Ap | -0,243 | 259 | <0,001 |

The obtained data suggest an increase in the popularity of search queries related to “migraine” and “headache” specifically during periods of decreased geomagnetic activity, and a corresponding decrease in the popularity of these queries during geomagnetic storms. Additionally, it is important to emphasize that patients with TTH are more sensitive to low geomagnetic activity than patients with migraine. This may indicate differences in the pathophysiological mechanisms of TTH and migraine, explaining the differences in sensitivity to external influences.

However, it is important to note that despite the relatively weak correlation between these changes, the obtained correlation coefficients (Table 2) are statistically significant. This suggests that fluctuations in the magnetic field, although they may act as triggers for migraines and tension-type headaches, are not the sole or dominant factors. Other factors may also influence these results, such as weather conditions, stress, changes in food consumption, or the influence of other external factors.

## Conclusion

The popularity of search queries for “migraine” and “headache” in the Google search engine can be a useful tool for identifying triggers of headaches. The obtained results demonstrate a negative correlation between the popularity of search queries for “migraine” and “headache” in Ukraine and changes in geomagnetic activity on Earth. During periods of decreased geomagnetic activity, concern among Google users in Ukraine regarding “migraine” and “headache” increases, and vice versa. Patients with TTH are more sensitive to low geomagnetic activity than patients with migraine. This may indicate differences in the pathogenetic mechanisms underlying these two types of primary headaches.

Overall, our study opens the door for further research in this direction and emphasizes the importance of understanding the impact of geomagnetic activity on human health.

## Data Availability

All data produced in the present study, including search query statistics and geomagnetic activity indices, are available upon reasonable request to the corresponding author.
All relevant data produced in the present work, including summary statistics and correlation results, are contained within the manuscript and its tables.
All data produced are available online at Google Trends and the Space Weather Prediction Center.

https://trends.google.com/

https://www.swpc.noaa.gov/

## Author Contributions

1. K. Yarovaya: contributed to the conceptualization of the study, data analysis, and interpretation of results, drafted and revised the manuscript.
2. I. Hnatiuk: assisted with data collection, performed statistical analyses, and contributed to manuscript revision.
3. Y. Solodovnikova: helped in the formulation of the research questions, analysis of the data, and drafting the manuscript.
4. A. Son: contributed to the methodology, literature review, and final manuscript review.

